# Potential of machine learning to predict early ischemic events after carotid endarterectomy or stenting: A comparison with surgeon predictions

**DOI:** 10.1101/2020.11.16.20231639

**Authors:** Kazuya Matsuo, Atsushi Fujita, Kohkichi Hosoda, Jun Tanaka, Taichiro Imahori, Taiji Ishii, Masaaki Kohta, Kazuhiro Tanaka, Yoichi Uozumi, Hidehito Kimura, Takashi Sasayama, Eiji Kohmura

## Abstract

**Background:** Carotid endarterectomy (CEA) and carotid artery stenting (CAS) are recommended for high stroke-risk patients with carotid artery stenosis to reduce ischemic events. However, we often face difficulty in determining the best treatment strategy.

**Objective:** We aimed to develop an accurate post-CEA/CAS outcome prediction model using machine learning that will serve as a basis for a new decision support tool for patient-specific treatment planning.

**Methods:** Retrospectively collected data from 165 consecutive patients with carotid stenosis underwent CEA or CAS were divided into training and test samples. The following five machine learning algorithms were tuned, and their predictive performance evaluated by comparison with surgeon predictions: an artificial neural network, logistic regression, support vector machine, random forest, and extreme gradient boosting (XGBoost). Seventeen clinical factors were introduced into the models. Outcome was defined as any ischemic stroke within 30 days after treatment including asymptomatic diffusion-weighted imaging abnormalities.

**Results:** The XGBoost model performed the best in the evaluation; its sensitivity, specificity, positive predictive value, and accuracy were 31.9%, 94.6%, 47.2%, and 86.2%, respectively. These statistical measures were comparable to those of surgeons. Internal carotid artery peak systolic velocity, low density lipoprotein cholesterol, and procedure (CEA or CAS) were the most contributing factors according to the XGBoost algorithm.

**Conclusion:** We were able to develop a post-procedural outcome prediction model comparable to surgeons in performance. The accurate outcome prediction model will make it possible to make a more appropriate patient-specific selection of CEA or CAS for the treatment of carotid artery stenosis.

## Introduction

Carotid artery stenosis is an important cause of ischemic stroke, which remains a major public health problem worldwide [26]. To reduce the risk of ischemic stroke, carotid endarterectomy (CEA) and carotid artery stenting (CAS) are recommended for patients at high stroke risk with carotid artery stenosis. Based on the evidence that there are no significant differences in long-term outcomes after CEA and CAS [1,10,23], there are general guidelines for patient selection for CEA and CAS [10,13,27]. However, we often face difficulty in determining the best treatment method. Therefore, it is necessary to develop a useful decision support tool for patient-specific treatment planning for carotid artery stenosis. Recently, the use of artificial intelligence (AI) has been increasing in medical field because of advances in technology including robust machine learning (ML) algorithms for successful prediction and diagnosis [14]. However, no studies have applied modern ML models in the carotid stenosis cohort.

In this study, we aimed to develop an accurate ML model for the prediction of ischemic events within 30 days after CEA or CAS with 17 clinical factors. The usefulness of the ML models was evaluated by comparing their predictions with those of surgeons. Because early periprocedural major and minor stroke is associated with long-term outcomes [18], our model to predict post-procedural ischemic events can serve as the basis for an effective decision support tool for patient-specific adaptation of CEA and CAS. Additionally, the relative importance of the clinical features was measured using an ML method.

## Materials and Methods

### Study population

We enrolled 170 consecutive cases of carotid stenosis treated with CEA or CAS at a single center in Japan between January 2013 and December 2018. Patient information was retrospectively collected from the hospital carotid stenosis database and anonymized before analysis. This retrospective study was approved by the institutional review board. Written informed consent was obtained from all patients before treatment. We excluded patients with arterial dissection and those who could not undergo MRI because of pacemaker implantation. The missing values were imputed by the k-nearest-neighbours (kNN) method [30]. We used the clinical data of patients until March 2018 as training data to optimize the hyperparameters and train the ML models, and used the data of more recent patients, from April to December 2018, as test data to evaluate the predictive performance of each model. The sample size was determined based on a previous study [8]. We conducted and reported this study in compliance with the transparent reporting of a multivariable prediction model for individual prognosis or diagnosis (TRIPOD) statement for multivariate prediction models [19].

### Treatments

Therapeutic approach was in accordance with the guidelines [13,27] based on stenosis degree assessed by NASCET criteria [7]. Basically, CAS was performed for CEA high-risk patients according to the inclusion criteria in the SAPPHIRE study [10]. The final treatment strategy was made by a multidisciplinary team. CEA was performed under general anaesthesia by three surgeons. Continuous neurophysiological monitoring was performed by neurophysiologists during surgery with a multimodality protocol involving electroencephalogram (EEG), median nerve somatosensory evoked potentials (SSEP), and bilateral regional cerebral oximetry (rSO2). Shunt placement was determined by the onset of alarm criteria for either EEG or SSEP, which was defined as a >50% decrease in amplitude [9]. CAS was performed under local anaesthesia by four surgeons. The rSO2 was monitored during the procedure. Each patient treated with CAS was administered 100 mg aspirin and 75 mg clopidogrel daily for at least 7 days before and 90 days after the CAS. The choice of stent and interventional strategy were determined by the interventional team.

### Clinical parameters

A total of 17 clinical parameters were used for the ML model development based on their known or expected influence on the outcome. These parameters consisted of age [28], pretreatment modified Rankin scale (mRS) [21], hypertension [21], diabetes mellitus [11,24,31], medical history of arteriosclerotic disease [5], serum low-density lipoprotein (LDL) cholesterol value (mg/dL) [24], internal carotid artery peak systolic velocity (ICA-PSV, cm/sec), symptomatic [24,28], crescendo transient ischemic attack (TIA) or stroke in evolution [11], previous neck irradiation [6], type III aorta [24], contralateral carotid occlusion [31], stenosis at a high position, mobile plaque, plaque ulceration [11,24], plaque with hyperintense signal on time-of-flight (TOF), and procedure (CEA or CAS) [4,18,20]. History of arteriosclerotic disease was defined as a history of acute coronary syndrome or peripheral artery disease requiring treatment. Crescendo TIA was defined as at least two similar TIAs in one week. Contralateral carotid occlusion, stenosis at a high position, and type III aorta were assessed using MRA, CTA, or angiography. Stenosis at a high position was defined as carotid stenosis that extends distally to the height of the vertebral body of C2. The ICA-PSV, mobile plaque, and plaque ulceration were assessed using echocardiogram. A plaque with hyperintense signal on TOF was defined as a plaque that appeared hyperintense on TOF-MRI compared with the signal of the adjacent sternocleidomastoid muscle.

In our preliminary study, we first created a prediction model with 22 clinical parameters which included gender, arterial fibrillation, estimated glomerular filtration rate, stenosis degree assessed by NASCET criteria, and the side of the lesion. We then find the effective parameters and narrowed it down to the 17 parameters with the highest predictive performance (data not shown).

### Outcomes

Outcome was defined as minor or major ischemic stroke including asymptomatic diffusion weighted imaging (DWI) hyperintense lesions within 30 days after CEA or CAS. Minor stroke was defined as a National Institutes of Health Stroke Scale score of less than 3, and major stroke was defined as a score of 3 or higher. Postprocedural MRI was performed the day after the procedure for CAS and less than one week afterwards for CEA. Additional MRI was performed if any neurological deficit was observed.

### Development of machine learning models

The following five ML models were applied: artificial neural network (ANN), logistic regression, random forest, support vector machine (SVM), and extreme gradient boosting (XGBoost) [3]. The logistic regression, random forest, and SVM were implemented using scikit-learn, which is a free ML library for Python. The ANN model was implemented using the Keras library with a TensorFlow backend. All the ML models were developed using Python version 3.7.7, Scikit-learn version 0.22.1, and TensorFlow version 2.2.0. First, we performed hyperparameter tuning of all models except for ANN using a grid-search algorithm with log-loss as the objective function on the training data. All numerical variables were standardized using centring and scaling. When applying grid-search, the value of the objective function was evaluated by stratified 5-fold cross-validation. The hyperparameters of the ANN model were hand-tuned using the holdout method on the training data with cross-entropy as the objective function. The base ANN model consisted of three dense layers with two dropout layers and two batch normalization layers (Supplemental table 1). After identifying the optimal hyperparameters for every model that minimize the log-loss value, we evaluated the predictive performance of each model using 10 times repeated stratified 5-fold cross-validation on the training data. The averages of the area under the receiver operating characteristic curve (ROC AUC), sensitivity, specificity, positive prediction value (PPV), and prediction accuracy were calculated through the cross-validation, and 95% confidence intervals were estimated. We also created and evaluated an ensemble model which comprises of the three models with the highest ROC AUC on the training data.

### Test of machine learning models

The data of 22 consecutive patients treated by CEA or CAS at the same institution from April to December 2018 were used as a test data. Their post-procedural outcomes were predicted using the trained ML models with the 17 factors. In this procedure, we used the standard bootstrap method as an additional internal validation technique. In this study, the number of resampling repetitions, which should be as large as possible to ensure the stability of the estimates, was set to 1000 repetitions. The bootstrap averages of sensitivity, specificity, PPV, and prediction accuracy were calculated, and 95% confidence intervals were estimated. Using the same 17 factors, four surgeons (board-certified neurosurgeons who had at least 10 years of experience) also predicted the post-procedural outcomes for each patient based on the test data within 10 minutes. When surgeons performed the outcome prediction test, to ensure that other information was never leaked, a paper test with information on only the 17 clinical factors for each patient was used. These surgeons were independent of the CEA and CAS performed during the designated study period. The average sensitivity, specificity, PPV, and prediction accuracy of the predictions of the four surgeons were compared with those of the ML models.

### Feature importance measurement

The relative importance of the clinical features was measured by the total gain of the XGBoost algorithm. The gain is the relative contribution of a feature to the model, calculated by taking each feature’s contribution to each tree in the gradient boosting decision tree model. Thus, the features with higher gain are more important for generating the prediction of the XGBoost model.

### Statistical Analysis

Descriptive and comparative statistics were used to describe clinical characteristics of the patients. We performed a statistical comparison between training and test groups using Welch’s t-test for numerical values, Fisher’s exact test for categorical variables, and the Mann–Whitney U test for pretreatment mRS. A 2-tailed probability value of 0.05 or lower was considered statistically significant. Statistical analysis was performed using EZR version 1.38.

## Results

### Study participants

The flow diagram of model development and validation is presented in Figure 1. Among the 170 patients with carotid stenosis, five (2.9%) were excluded. Thus, the data of a total of 165 patients with carotid stenosis were included for analysis and separated into training and test data. The baseline characteristics before missing value imputation are shown in Table 1. There were 36 (22%) patients over 80 years of age, and 127 (77%) patients in good condition (mRS 0-1). Severe carotid stenosis, which was defined as an ICA-PSV of more than 200 cm/s, was observed in 115 (70%) patients. CEA and CAS were performed on 95 (58%) and 70 (42%) patients, respectively. The outcome was observed in 45 (27%) patients. Major stroke, minor stroke, and asymptomatic DWI hyperintense lesions were diagnosed in 3 (1.8%), 3 (1.8%), and 39 (24%) patients, respectively. All missing values were imputed using the kNN method. No significant difference was found in the comparison before and after imputation of the missing values (data not shown). Although age and follow-up duration were significantly older and shorter, respectively, in the test data, there was no significant difference between the training and test data for the other factors that were used for analysis after imputation (Table 1). The test data tended to have more CAS and fewer outcomes, but there were no significant differences.

**Table 1.** Patient characteristics

| Variable | Before imputation | After imputation |  | *p* value |
| --- | --- | --- | --- | --- |
|  | Total (n=165) | Training data (n=143) | Test data (n=22) |  |
| Age, year, mean (SD) | 74.2 (7.61) | 73.8 (7.82) | 76.7 (5.62) | 0.04 |
| Male sex, n (%) | 141 (85) | 122 (85) | 19 (86) | 1 |
| pre-treatment mRS, median [IQR] | 0 [0-1] | 0 [0-1] | 0.5 [0-1.75] | 0.55 |
| LDL-cholesterol, mg/dL, mean (SD) | 90.0 (29.5) | 90.4 (30.1) | 87.1 (23.3) | 0.56 |
| Prior medical histories, n (%) |  |  |  |  |
| Hypertension | 136 (82) | 118 (83) | 18 (82) | 1 |
| Diabetes mellitus | 58 (35) | 50 (35) | 8 (36) | 1 |
| Acute coronary syndrome | 53 (32) | 42 (29) | 11 (50) | 0.08 |
| Peripheral artery disease | 28 (17) | 24 (17) | 4 (18) | 0.77 |
| Anatomical and pathophysiological features |  |  |  |  |
| Contralateral occlusion, n (%) | 14 (8.5) | 11 (7.7) | 3 (14) | 0.40 |
| Stenosis at a high position, n (%) | 13 (7.9) | 12 (8.4) | 1 (4.5) | 1 |
| Type III Aorta, n (%) | 64 (39) | 56 (39) | 8 (36) | 1 |
| ICA-PSV, cm/sec, mean (SD) | 276 (130) | 272 (126) | 291 (147) | 0.56 |
| Mobile plaque, n (%) | 19 (12) | 15 (10) | 4 (18) | 0.29 |
| Plaque ulceration, n (%) | 39 (24) | 36 (25) | 3 (14) | 0.29 |
| plaque with hyperintense signal on TOF, n (%) | 61 (37) | 54 (38) | 7 (32) | 0.64 |
| Previous neck irradiation, n (%) | 15 (9.1) | 13 (9.1) | 2 (9.1) | 1 |
| Symptomatic, n (%) | 64 (39) | 56 (39) | 8 (36) | 1 |
| Crescendo TIA or stroke in evolution, n (%) | 10 (6.1) | 8 (5.6) | 2 (9.1) | 0.62 |
| Treatment, CEA, n (%) | 95 (58) | 85 (59) | 10 (45) | 0.25 |
| Outcome, n (%) |  |  |  |  |
| Ischemic stroke within 30 days | 45 (27) | 42 (29) | 3 (14) | 0.13 |
| Major ischemic stroke | 3 (1.8) | 2 (1.4) | 1 (4.5) | 0.36 |
| Minor ischemic stroke | 3 (1.8) | 3 (2.1) | 0 (0) | 1 |
| Asymptomatic DWI hyperintense lesions | 39 (24) | 37 (26) | 2 (9.1) | 0.11 |
| Follow-up duration, days, mean (SD) | 833 (565) | 921 (555) | 259 (259) | <.0001 |

CEA = carotid endarterectomy; DWI = diffusion weighted imaging; IQR = interquartile range; LDL = low density lipoprotein; mRS = modified Rankin scale; ICA-PSV = internal carotid artery-peak systolic velocity; TIA = transient ischemic attack; TOF = time-of-flight.

**Fig. 1.**
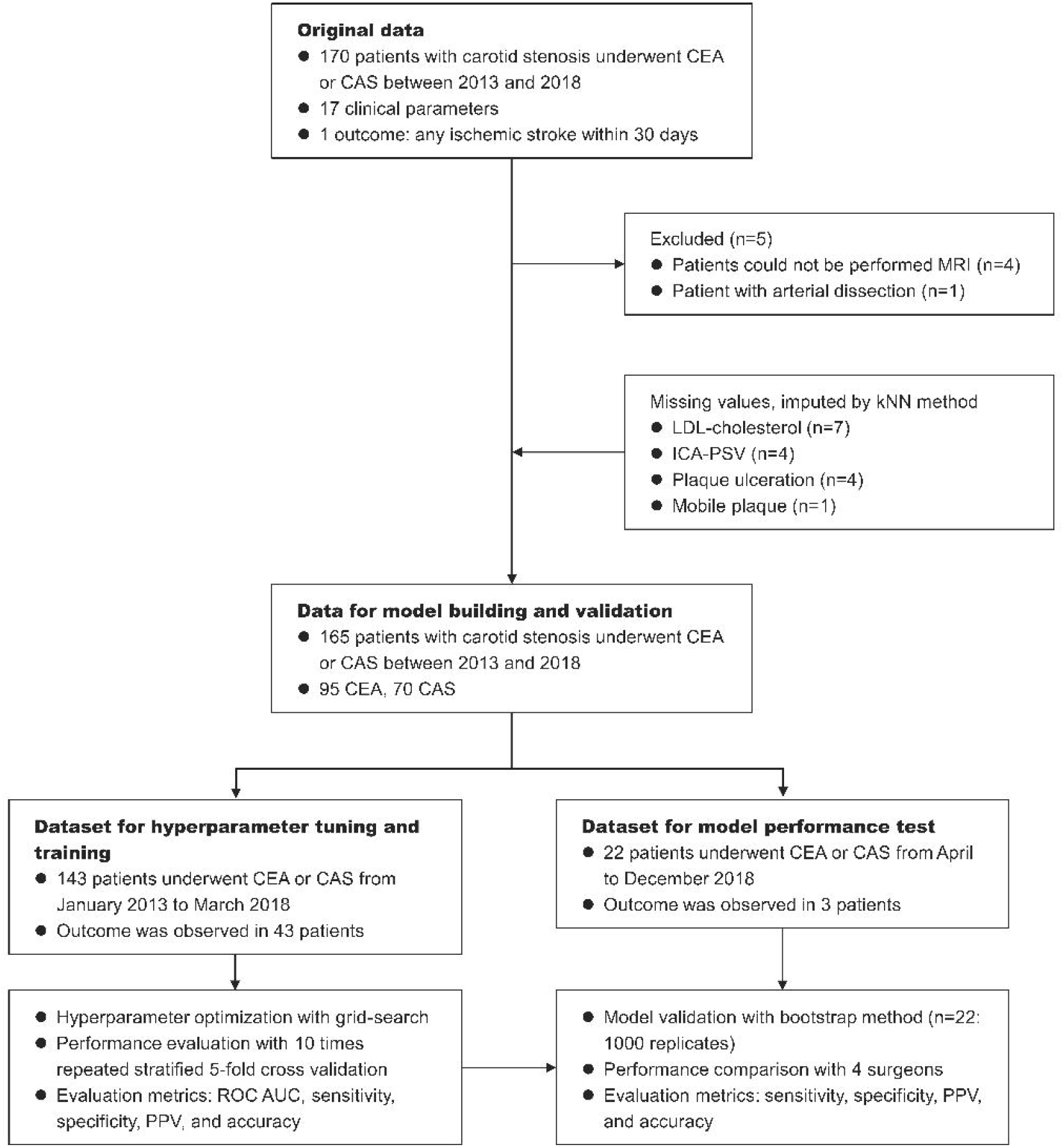
Flow diagram describing the general framework of the study. Models were built using the training dataset. The test dataset was used for measuring the predictive performance and comparison with the surgeons. CAS, carotid artery stenting; CEA, carotid endarterectomy; ICA-PSV, internal carotid artery peak systolic velocity; kNN, k-nearest-neighbours; LDL, low density lipoprotein; PPV, positive predictive value

The differences in patient characteristics between the CEA and CAS groups were shown in Supplemental table 1. The patients with hypertension, plaque ulceration, plaque with hyperintense signal on TOF, and symptomatic stenosis were significantly more common in the CEA group. Asymptomatic DWI hyperintense lesion was significantly increased in CAS group. Other variables did not differ between the CEA and CAS groups.

### Prediction of post-procedural ischemic events on the training dataset

To evaluate the predictive performance of five ML models for post CEA/CAS outcomes, 10 times repeated stratified 5-fold cross-validation was first performed on the 143-patient training data. The prediction results showed that the ROC AUC of XGBoost was highest at 0.719, sensitivity of SVM was highest at 36.2%, and the specificity, PPV, and accuracy of random forest were highest at 98.3%, 78.9%, and 75.4%, respectively (Table 2). Then, an ensemble model of the logistic regression, XGBoost, and ANN models, which were the three most highest ROC AUC models, was also created and evaluated. It yielded a ROC AUC of 0.739, sensitivity of 15.1%, specificity of 97.4%, PPV of 75.1%, and accuracy of 72.7%. Thus, the ensemble model obtained the highest ROC AUC on the training data.

**Table 2.** Prediction results on the training dataset evaluated by repeated 5-fold cross validation and sorted by ROC AUC

| **Model** | **ROC AUC**<br>mean (95%CI) | **Sensitivity (%)**<br>mean (95%CI) | **Specificity (%)**<br>mean (95%CI) | **PPV (%)**<br>mean (95%CI) | **Accuracy (%)**<br>mean (95%CI) |
| --- | --- | --- | --- | --- | --- |
| Ensemble model * | 0.739<br>(0.714 - 0.764) | 15.1<br>(11.3 – 18.8) | 97.4<br>(96.2 – 98.6) | 75.1<br>(65.6 – 84.7) | 72.7<br>(71.3 – 74.0) |
| XGBoost | 0.719<br>(0.692 - 0.746) | 14.6<br>(10.3 – 19.0) | 94.9<br>(93.1 – 96.7) | 54.5<br>(44.6 – 64.3) | 70.8<br>(69.1 – 72.4) |
| Logistic regression | 0.702<br>(0.671 - 0.732) | 26.5<br>(22.4 – 30.5) | 95.5<br>(94.2 – 96.8) | 71.2<br>(62.7 – 79.7) | 74.8<br>(73.1 – 76.4) |
| Neural network | 0.692<br>(0.659 - 0.726) | 29.0<br>(23.8 – 34.1) | 91.2<br>(89.1 – 93.3) | 61.3<br>(54.3 – 68.3) | 72.5<br>(70.8 – 74.3) |
| Random forest | 0.683<br>(0.653 - 0.712) | 22.0<br>(18.0 – 26.0) | 98.3<br>(97.4 – 99.2) | 78.9<br>(69.2 – 88.7) | 75.4<br>(74.0 – 76.8) |
| SVM | 0.680<br>(0.650 - 0.711) | 36.2<br>(32.2 – 40.3) | 85.3<br>(83.5 – 87.1) | 51.7<br>(46.6 – 56.8) | 70.6<br>(68.8 – 72.4) |

\* Ensemble model is created by using XGBoost, neural network, and logistic regression, which are the three most highest ROC AUC models.
6 ROC AUC = area under the receiver operating characteristic curve; PPV = positive predictive 7 value; SVM = support vector machine.

### Prediction of post-procedural ischemic events on the test dataset and comparison with surgeons

Next, to confirm the post-CEA/CAS outcome prediction performances of the six ML models including the ensemble model, these models were further evaluated on the 22-patient test data with the bootstrap method. The results are shown in Table 3. XGBoost achieved the highest PPV, and accuracy scores, which were 47.2% and 86.2%, respectively. The highest sensitivity was 34.0%, which was achieved by random forest, SVM, and logistic regression. The highest specificity was 95.4%, which was achieved by ANN. The ensemble model, which obtained the highest ROC AUC on the training data, did not perform so well on the test data.

**Table 3.** Prediction results on the test dataset evaluated using the bootstrap technique and sorted by accuracy.

| **Model** | **Sensitivity (%)**<br>mean (95%CI) | **Specificity (%)**<br>mean (95%CI) | **PPV (%)**<br>mean (95%CI) | **Accuracy (%)**<br>mean (95%CI) |
| --- | --- | --- | --- | --- |
| XGBoost | 31.9<br>(30.0 – 33.8) | 94.6<br>(94.2 – 94.9) | 47.2<br>(44.9 – 49.6) | 86.2<br>(85.7 – 86.6) |
| Neural network | 4.1<br>(3.3 – 4.8) | 95.4<br>(95.3 – 95.5) | 12.4<br>(10.4 – 14.4) | 83.3<br>(82.9 – 83.8) |
| Ensemble model * | 31.8<br>(29.9 – 33.7) | 89.6<br>(89.2 – 90.0) | 32.0<br>(30.0 – 33.9) | 82.0<br>(81.5 – 82.5) |
| Random forest | 34.0<br>(32.1 – 35.9) | 84.3<br>(83.8 – 84.8) | 25.1<br>(23.6 – 26.6) | 77.7<br>(77.1 – 78.2) |
| SVM | 34.0<br>(32.1 – 35.9) | 79.1<br>(78.6 – 79.7) | 19.9<br>(18.7 – 21.1) | 73.2<br>(72.6 – 73.7) |
| Logistic regression | 34.0<br>(32.1 – 35.9] | 79.0<br>(78.4 – 79.6) | 20.2<br>(18.9 – 21.5) | 73.0<br>(72.4 – 73.6) |
| Surgeons ** | 41.7<br>(0 – 92.4) | 75.0<br>(60.7 – 89.3) | 20.1<br>(0 – 42.8) | 70.5<br>(57.9 – 83.0) |

\* Ensemble model is created by using XGBoost, neural network, and logistic regression, which are the three most highest ROC AUC models on the training dataset. \*\* The average of 4 surgeon's prediction results with 95% confidence interval. PPV = positive predictive value; SVM = support vector machine.

The average of the outcome predictions made by four surgeons had a sensitivity of 41.7%, specificity of 75.0%, PPV of 20.1%, and accuracy of 70.5%. Therefore, all of the current ML models trained with 143 cases and 17 factors outperformed the surgeons’ predictions in terms of specificity and accuracy. However, surgeons showed a higher predictive sensitivity than the current ML models. A statistical analysis was not performed on these results because of the small sample size. The optimized hyperparameters of these models are listed in Supplemental table 2.

### Importance values of the clinical factors

The feature importances were measured using a function of the XGBoost algorithm, which obtained the best predictive performance on the test data. The results reveal that ICA-PSV, serum LDL-cholesterol value, and procedure (CEA or CAS) are the most important in this order (Figure 2).

**Fig. 2.**
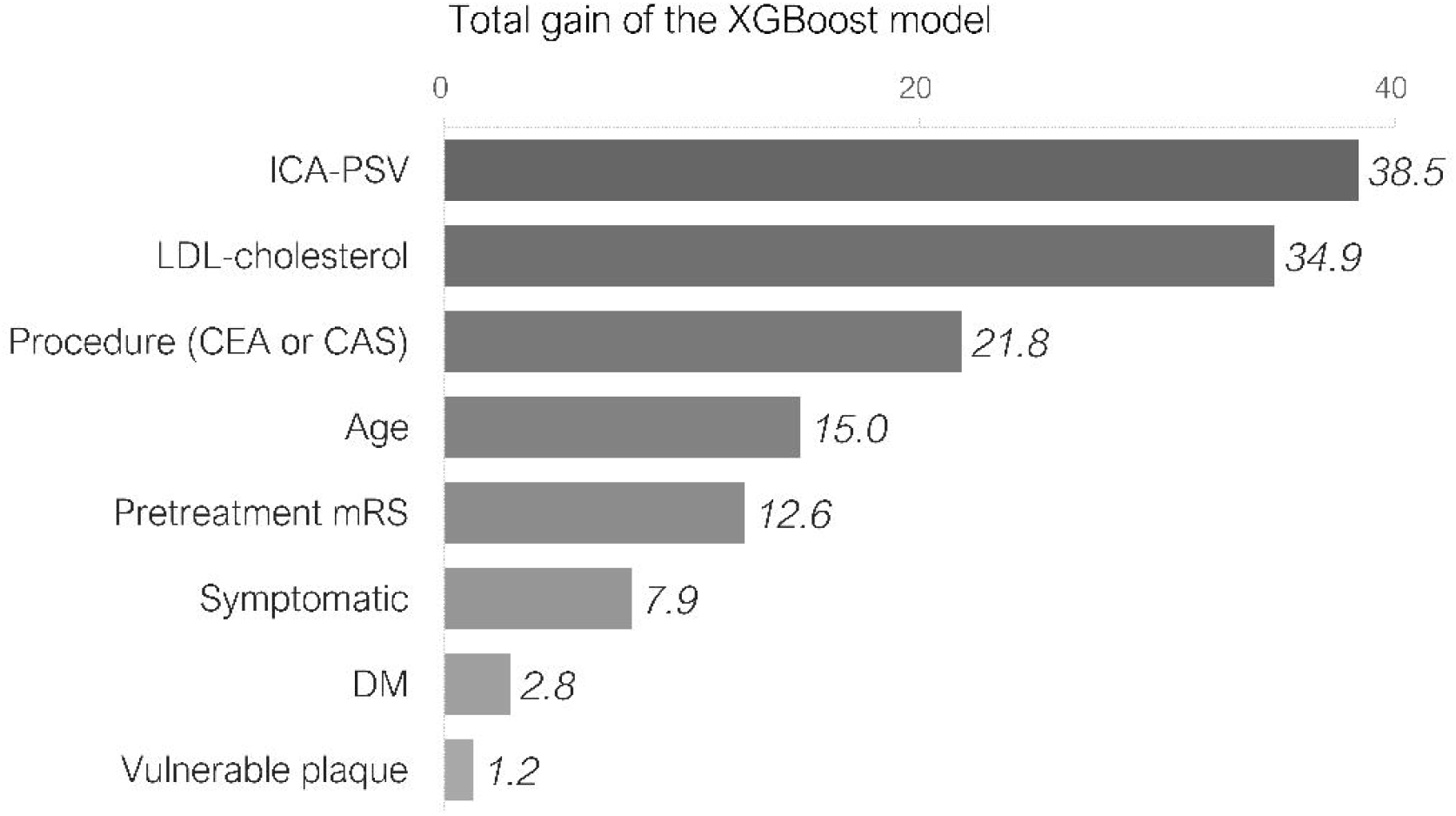
Importance values of the clinical factors measured using the total gain of the XGBoost algorithm. CAS, carotid artery stenting; CEA carotid endarterectomy; DM, diabetes mellitus; ICA-PSV, internal carotid artery peak systolic velocity; LDL, low density lipoprotein; mRS, modified Rankin scale

## Discussion

In this study, we identified two notable findings. First, using an appropriate model construction process with effective clinical factors, we were able to develop a post-CEA/CAS outcome prediction model that is comparable to surgeons in terms of sensitivity, specificity, PPV, and accuracy, although our models were developed with a relatively small number of patients. Second, in our validation process, the XGBoost model had the highest predictive performance, and the factors that contributed most to the accurate model were ICA-PSV, serum LDL-cholesterol value, and procedure (CEA or CAS).

### ML and ICAS

We were able to develop a post-CEA/CAS outcome prediction model that is comparable to surgeons, even with a relatively small training sample size. This is the first study to predict the outcome after treatment of carotid artery stenosis using ML models. Our model makes it possible to preoperatively calculate the post-CEA/CAS stroke risk as a concrete numeric value; for example, 10% with CEA and 15% with CAS for postprocedural stroke risk. This function can help surgeons to determine whether the patient suitable for CEA or CAS. However, a problem of the current models is that sensitivity is relatively low. To increase the prediction sensitivity, we tuned the models with sensitivity as the objective function during model development. However, such sensitivity-oriented models showed a considerable decrease in accuracy, although sensitivity increased slightly (data not shown). Therefore, these sensitivity-oriented models were not adopted this time. In this study, we did not perform statistical analyses on differences of predictive performance between ML models and surgeons because of the small test sample size. Therefore, we did not determine the statistical significance of ML models. However, our prediction models could be a milestone in the development of new decision support tool for treatment choice in carotid stenosis, because as the training data continues to grow in size, we can expect the prediction performance of the models to be higher than the present study. To further improve the predictive performance of ML models, it would be necessary to use big data and more important contributing factors.

In this study, the incidence rate of postprocedural ischemic events was 27%, which was similar to the results of previous studies [29]. Most of these postprocedural ischemic events were asymptomatic DWI small lesions. Although such asymptomatic DWI lesions are sometimes underestimated in clinical practice, it was reported that the volume of postoperative DWI lesions could correlate with cognitive deterioration at 6 months after CEA or CAS [36]. Accordingly, even if the lesion is small, we should try to avoid the postoperative DWI lesions. In general, MRI DWI can detect ischemic changes for at least 2 weeks after a stroke onset, and the postoperative DWI lesions have been considered to be derived from cerebral microembolism during CAS or CEA [25]. Thus, the difference in the timing of postoperative MRI between the CEA and CAS groups should not have a significant impact on our results.

The number of studies using ML to explore stroke and neurosurgical diseases has increased rapidly over the past decade. The most frequently applied algorithms are ANN, logistic regression, random forest, and SVM [2,12,32], which suggests that our model selection is appropriate. In our study, the predictive performance of the XGBoost was better than those of the ANN, logistic regression, SVM, and random forest. Although the superior performance of the GBDT model has been shown in many data-science contests in recent years [33], there is still very little research on neurosurgery or stroke using the GBDT model [22,33]. The ensemble model, which had the highest ROC AUC on our training data, did not perform so well on the test data, probably because of overfitting to the training data.

### Feature Importance

XGBoost, which showed the best predictive performance, identified ICA-PSV, serum LDL-cholesterol value, and procedure (CEA or CAS), as the most effective factors. The procedure type, CEA or CAS, has previously been reported as a potential predictor of periprocedural stroke [4,18,20,21]. Thus, it seems reasonable that the procedure type is the third most important factor in this study. However, no studies have suggested that ICA-PSV or serum LDL-cholesterol value are associated to postprocedural outcome. Although ICA-PSV should reflect the degree of stenosis, many studies reported that the degree of stenosis is not associated with postprocedural outcome [24,28]. ICA-PSV might be used in our model as a valid predictor in combination with other clinical factors. In a study investigating the components of emboli captured by the filter-protection device during CAS, more postoperative DWI high intensity lesions were observed in patients whose main component of the emboli was cholesterol [17]. It may be inferred from such results that serum LDL-cholesterol might be related to post-procedural outcome because it should be possible for a higher serum LDL-cholesterol value to leads to a higher cholesterol content of the plaque, which produces emboli. In this study, the presence and intensity of lipid-lowering therapies, such as statins, were not evaluated. How LDL-cholesterol levels and lipid-lowering therapies are associated with clinical outcome need to be further investigated in future studies.

### Comparison of clinician and AI performance

Many studies have compared clinicians and AI with image interpretation or diagnostic performance and have shown that ML models are equivalent to or superior to specialists [16,34]. However, there were very few studies that compared the predictive performance of clinicians and AI as in this study [15]. The sensitivity of the surgeons’ prediction was higher than our ML models, presumably because the surgeons have experience of more than 143 patients, which comprise the dataset that our ML models learned. Therefore, instead of a difference in predictive ability, the number of experienced and learned cases might have influenced the results. The advantage of an ML model is that if only the question and answer are provided correctly, it can learn an enormous number of cases that one surgeon would not be able to experience. Therefore, in future, as the number of training cases increases, the predictive performance of the ML model would further improve.

### Limitations

One of the main limitations of our study is the small sample size. Although a larger sample size would be needed for more accurate ML models, a previous report suggested that 80–560 samples are required for ML algorithms excluding deep neural networks, and the required sample size depends on the dataset and sampling method [19]. Furthermore, a systematic review of AI in neurosurgery has shown that the median number of patients in each study was 120 [2]. Thus, the sample size might not be insufficient for our ML models (excepting the ANN model). Second, several potentially important clinical parameters, such as tandem stenotic lesions or other inappropriate anatomical features for the procedure, were not considered in this study. Third, optimizing hyperparameters for neural network models is generally difficult because they have many hyperparameters that need to be adjusted. In addition to hyperparameters, the neural network architecture should be optimized for better performance [35]. In this study, because the ANN was hand-tuned by multiple trial-and-error sessions, a more effective hyperparameter set might be found by other, more sophisticated optimization methods. Finally, because the dataset was collected retrospectively from a single institution, it is prone to selection bias, and our ML models may not be applicable to other institutions where different treatment strategies or patient demographics might exist. Although internal validation was applied with repeated cross-validation and bootstrap methods, further external validation is necessary in another setting that differs in time or place to validate the performance of our prognostic models.

## Conclusion

We developed a post-CEA/CAS outcome prediction model with a performance comparable to that of surgeons. The XGBoost model showed the best predictive performance which achieved more than 85% accuracy, and the most contributing factors were ICA-PSV, serum LDL-cholesterol value, and procedure (CEA or CAS). Our model can help surgeons to determine whether the patient suitable for CEA or CAS based on the calculated probability estimates for postprocedural ischemic event. Larger datasets and analysis of potential prognostic factors would be necessary to further improve the predictive performance of the ML models.

## Data Availability

The patient data that support the findings of this study are available from the corresponding author upon reasonable request. A subset of the program code generated for this study is available at GitHub.

https://gist.github.com/kkmatsuo/e77e78a0346280d5570829164760132f

## Declarations

### Availability of data and material (data transparency)

The patient data that support the findings of this study are available from the corresponding author upon reasonable request.

### Code availability (software application or custom code)

A subset of the program code generated for this study is available at GitHub and can be accessed at https://gist.github.com/kkmatsuo/e77e78a0346280d5570829164760132f.

### Ethics approval (include appropriate approvals or waivers)

This retrospective study was approved by the Ethical Committee of Kobe University Graduate School of Medicine (approval no. B200444).

### Consent to participate (include appropriate statements)

Written informed consent was obtained from all patients before treatment.

### Consent for publication (include appropriate statements)

Not applicable.

### Authors’ contributions

Kazuya Matsuo conceptualized the experimental design, analyzed the data, developed the machine learning model, and wrote the manuscript. Atsushi Fujita contributed to study design, interpreted the results, and critically revised the manuscript. Kohkichi Hosoda provided the background data for the study, interpreted the results, and reviewed the manuscript. Jun Tanaka carried out the data acquisition, conducted the analyses, and reviewed the manuscript. Taichiro Imahori carried out the data acquisition and reviewed the manuscript. Taiji Ishii carried out the data acquisition and reviewed the manuscript. Masaaki Kohta participated in the study design, interpreted the results, and reviewed the manuscript. Kazuhiro Tanaka carried out the data acquisition, conducted the analyses, and interpreted the results. Yoichi Uozumi conducted the analyses and interpreted the results. Hidehito Kimura carried out the data acquisition and interpreted the results. Takashi Sasayama supervised the whole study and critically revised the manuscript. Eiji Kohmura supervised the whole study, contributed to study design, and critically revised the manuscript. All authors read and approved the final manuscript.

## Supplementary Information

**Supplemental Table 1** Patient characteristics of CEA group and CAS group.

**Supplemental Table 2** Optimized hyperparameters of five machine learning models.

