## Supplemental table1,2 for "Potential of machine learning to predict early ischemic events after carotid endarterectomy or stenting: A comparison with surgeon predictions"

**Article title**

**Journal name**

Neurosurgical Review

**Author names**

Kazuya Matsuo, Atsushi Fujita, Kohkichi Hosoda, Jun Tanaka, Taichiro Imahori, Taiji Ishii, Masaaki Kohta, Kazuhiro Tanaka, Yoichi Uozumi, Hidehito Kimura, Takashi Sasayama, Eiji Kohmura.

**Affiliation and e-mail address of the corresponding author**

Kazuya Matsuo, MD, PhD, Department of Neurosurgery, Kobe University Graduate School of Medicine, Kobe, Japan.

Supplemental Table 1. Patient characteristics of CEA group and CAS group.

| Variable | CEA group (n=95) | CAS group (n=70) | *p* value |
|---|---|---|---|
| Age, year, mean (SD) | 74.0 (8.22) | 74.5 (6.74) | 0.70 |
| pre-treatment mRS, median [IQR] | 0 [0-1] | 0 [0-2] | 0.41 |
| LDL-cholesterol, mg/dL, mean (SD) | 89.7 (32.7) | 90.7 (23.9) | 0.83 |
| Prior medical histories, n (%) | | | |
| Hypertension | 84 (88) | 52 (74) | 0.02 |
| Diabetes mellitus | 30 (32) | 28 (40) | 0.32 |
| Arteriosclerotic disease | 36 (38) | 35 (50) | 0.15 |
| Anatomical and pathophysiological features | | | |
| Contralateral occlusion, n (%) | 9 (9.5) | 5 (7.1) | 0.78 |
| Stenosis at a high position, n (%) | 8 (8.4) | 5 (7.1) | 1 |
| Type III Aorta, n (%) | 33 (35) | 31 (44) | 0.26 |
| ICA-PSV, cm/sec, mean (SD) | 259 (125) | 294 (132) | 0.09 |
| Mobile plaque, n (%) | 14 (15) | 5 (7.1) | 0.15 |
| Plaque ulceration, n (%) | 33 (35) | 6 (8.6) | <0.0001 |
| Plaque with hyperintense signal on TOF, n (%) | 43 (45) | 18 (26) | 0.01 |
| Previous neck irradiation, n (%) | 7 (7.4) | 8 (11) | 0.42 |
| Symptomatic, n (%) | 45 (47) | 19 (27) | 0.01 |
| Crescendo TIA or stroke in evolution, n (%) | 3 (3.2) | 7 (10) | 0.10 |
| Outcome, n (%) | | | |
| Ischemic stroke within 30 days | 20 (21) | 26 (37) | 0.03 |
| Major ischemic stroke | 3 (3.2) | 0 (0) | 0.27 |
| Minor ischemic stroke | 0 (0) | 3 (4.3) | 0.07 |
| Asymptomatic DWI hyperintense lesions | 17 (18) | 23 (33) | 0.04 |

CAS = carotid artery stenting; CEA = carotid endarterectomy; DWI = diffusion weighted imaging; IQR = interquartile range; LDL = low density lipoprotein; mRS = modified Rankin scale; ICA-PSV = internal carotid artery-peak systolic velocity; TIA = transient ischemic attack; TOF = time-of-flight.

Supplemental table 2. Optimized hyperparameters of five machine learning models

| **Model** | **Hyperparameters** |
|---|---|
| XGBoost | learning_rate = 0.3<br>n_estimators = 360<br>max_depth = 1<br>min_child_weight = 6.5<br>gamma = 0.95<br>subsample = 0.65<br>colsample_bytree = 0.65<br>objective = 'binary:logistic'<br>reg_alpha = 0.001<br>reg_lambda = 0.1<br>max_delta_step = 1<br>early_stopping_rounds = 20 |
| Random forest | criterion = 'entropy'<br>max_depth = 4<br>max_features = 4<br>min_samples_leaf = 0.0001<br>min_samples_split = 0.0001<br>n_estimators = 40 |
| Logistic regression | penalty = 'l2'<br>C = 0.05<br>solver = 'saga' |
| SVM | kernel = 'linear'<br>C = 2.5<br>coef0 = 0<br>degree = 1<br>gamma = 0.0001<br>probability = True |

| | |
|---|---|
| Neural Network | models.Sequential()<br>model.add(layers.Dense(16, activation ='relu', input_shape=(17,)))<br>model.add(layers.Dropout(0.2))<br>model.add(layers.BatchNormalization())<br>model.add(layers.Dense(16, activation ='relu'))<br>model.add(layers.Dropout(0.2))<br>model.add(layers.BatchNormalization())<br>model.add(layers.Dense(1, activation ='sigmoid'))<br><br>EarlyStopping (patience=20, restore_best_weights=True)<br>optimizer=Adam (lr=0.01)<br>batch_size=8<br>epochs=41 |

SVM = support vector machine

* A subset of the program code generated for this study is available at GitHub and can be accessed at [BLINDED FOR REVIEW].
